# Use of cytokine-induced killer cell therapy in colorectal cancer patients: a systematic review and meta-analysis

**DOI:** 10.1101/2023.01.05.22283441

**Authors:** Celine Man Ying Li, Yoko Tomita, Bimala Dhakal, Runhao Li, Jun Li, Paul Drew, Timothy Price, Eric Smith, Guy J. Maddern, Kevin Fenix

## Abstract

**Background:** The number of clinical studies evaluating the benefit of cytokine-induced killer cell (CIK) therapy, an adoptive immunotherapy, for colorectal cancer (CRC) are increasing. In many of these trials CIK therapy was co-administered with conventional cancer therapy. The aim of this review is to systematically assess the available literature, in which the majority were only in Chinese, on CIK therapy for the management of CRC using meta-analysis, and to identify parameters associated with successful CIK therapy implementation.

**Methods:** Prospective and retrospective clinical studies which compared CIK therapy to non-CIK therapy in CRC patients were searched for electronically on MEDLINE, Embase, CNKI and Wanfang Data databases. The clinical endpoints of overall survival (OS), progression-free survival (PFS), OS and PFS rates, overall response rate (ORR) and toxicity were meta-analysed using hazard (HR) and relative ratios (RR), and subgroup analyses were performed using Chi-square (Chi^2^) test and I-square (I^2^) statistics for study design, disease stage, co-therapy type, and timing of administration.

**Results:** In total, 70 studies involving 6,743 patients were analysed. CIK therapy was favoured over non-CIK therapy for OS (HR=0.59, 95% CI: 0.53-0.65), PFS (HR=0.55, 95% CI: 0.47-0.63), and ORR (RR=0.65, 95% CI: 0.57-0.74) without increasing toxicity (HR=0.59, 95% CI 0.16-2.25). Subgroup analyses on OS and PFS by study design (randomised versus non-randomised study design), disease stage (Stage I-III versus Stage IV), co-treatment with dendritic cells (CIK versus DC-CIK therapy), or timing of therapy administration (concurrent versus sequential with co-administered anti-cancer therapy) also showed that the clinical benefit of CIK therapy was robust in any subgroup analysis. Furthermore, co-treatment with dendritic cells did not improve clinical outcomes over CIK therapy alone.

**Conclusions:** Compared with standard therapy, patients who received additional CIK cell therapy had favourable outcomes without increased toxicity, warranting further investigation into CIK therapy for the treatment of CRC.

**Key Message:** *What is already known on this topic:* Cytokine-induced killer cell (CIK) therapy is an adoptive immunotherapy used to treat both solid and haematological cancers for over 20 years. It is predominantly used in China, with multiple studies reporting benefit in colorectal cancer (CRC) patients. Despite this, CIK therapy treatment regimens are not widely used, possibly due in part to the majority of the literature about CIK therapy in CRC being reported in Chinese. Further, CIK therapy is commonly combined with other therapies but it is currently not known if there is a specific combination or treatment regimen that is optimal for CRC. What this study adds
We report the most comprehensive systematic review to date of CIK therapy for CRC patients, combining both Chinese and English language reports. Patients with CRC who received additional CIK therapy had better survival outcomes than with standard therapy alone. We also showed that the addition of dendritic cells (DC) to CIK therapy, common for CRC treatment, did not provide any clinical benefit over CIK therapy alone, and that CIK therapy is effective whether given concurrently or sequentially to standard treatment regimens.
How this study might affect research, practice, or policy
Our systematic review of Chinese and English publications shows that CRC patients benefit from the addition of CIK therapy to standard treatment protocols and warrants further international studies.

## Introduction

Colorectal cancer (CRC) is the second leading cause of cancer-related death worldwide^1^. Patients with locally advanced CRC, including regional lymph node metastases, have a 5-year survival of 75%, which reduces to 15% if there are distant metastases^2^. Survival outcomes for locally advanced and metastatic CRC have steadily improved due to advancements in surgical techniques, peri-operative care and therapeutic options. However, tumour recurrence and therapy resistance remain a challenge, creating the need for new treatment options^3^.

During the last decade immunotherapy has revolutionised cancer treatment, with clinical efficacy established for multiple solid and haematological cancers^4^. Immune checkpoint inhibitors have provided significant clinical benefit, particularly in solid cancers with a high tumour mutation burden^5^. In advanced CRC cases they have become the standard of care for high microsatellite instability (MSI-high)/mismatch repair deficient (dMMR) tumours^6,7^. Adoptive immunotherapy involves the administration of immune cells expanded and modified in *ex vivo* culture. Most treatments have focused on chimeric antigen receptor T (CAR-T) cell therapy. However, other technologies including dendritic cell (DC) therapy, natural killer (NK) cell therapy, and cytokine-induced killer cell (CIK) therapy are being studied. Unlike immune checkpoint inhibitors, none of the adaptive immunotherapy products are Food and Drug Administration (FDA) approved for CRC treatment^8^.

CIK therapy is an autologous, adoptive immunotherapy generated by expanding a heterogeneous population of immune effector cells from peripheral blood mononuclear cells (PBMC)^9^. The cell therapy product contains conventional T cells (CD3+CD56-), natural killer (NK)-like T cells (CD3+CD56+) and NK cells (CD3-CD56+)^10^. NK-like T cells are considered the main effector cells in CIK therapy, being able to recognise tumour cells in a major-histocompatibility complex (MHC)-class I unrestricted manner^11,12^. Hence, guidelines for CIK therapy patient transfusion require that the cell therapy product contain at least 40% NK-like T cells^13^. While CIK therapy is normally combined with conventional chemotherapy, multiple trials which combine CIK therapy with other immunotherapies are being investigated. One of the more popular combinations is combining CIK therapy with autologous DC therapy (DC-CIK therapy) with reports suggesting an improvement in anti-tumour activity^14^. China has been a leader in CIK therapy trials for multiple solid tumours, and CIK therapy is commonly provided for CRC treatment in some Chinese hospitals^15,16^.

To date there is a plethora of publications of varying study quality examining the clinical benefit of CIK therapy for CRC. The latest systematic review investigating the clinical efficacy of CIK therapy with chemotherapy in CRC patients was published in 2017^16^. Since then more studies have been published which support its clinical benefit^17-19,31^, warranting an updated systematic review to consolidate the evidence for CIK therapy in CRC management. Many of the reports originate in China and are written in Chinese. The objective of this work, therefore, is to systematically assess by meta-analysis the available literature on CIK therapy for the management of CRC, written in either English or Chinese. It includes both prospective and retrospective studies, and also analysed the benefit of parameters commonly modified in trials such as the addition of dendritic cells (DC-CIK therapy) or chemotherapy regimens.

## Methods

This systematic review and meta-analysis was performed according to the Preferred Reporting Items for Systematic Reviews and Meta-analysis statement^20^.

### Study selection and search

Studies which compared efficacy of CIK therapy, with or without another anti-cancer treatment, to no treatment or non-CIK anti-cancer treatment in adult patients with CRC diagnosis were identified MEDLINE, Embase, China National Knowledge Infrastructure (CNKI) and Wanfang Data databases. CNKI and Wanfang Data were included as there were multiple studies published in Chinese alone, which were not registered with Embase or MEDLINE. The search strategy for Embase and MEDLINE is described in Supplementary Tables S1 and S2, respectively. For CNKI and Wanfang Data, the following search keywords were used; “cytokine-induced killer cells”, “CIK”, “rectal cancer”, “colorectal cancer”, “colon cancer”, and “clinical trials”. No limits were placed on the language in which studies were published and the final search was performed in July 2022. Both prospective and retrospective studies with a parallel-arm design were considered, and the CIK therapy arm included patients who received CIK or DC-CIK (CIK/DC-CIK) therapy. Studies that did not report efficacy endpoints were excluded from this systematic review.

### Data extraction and quality assessment

Data collection was performed independently by two authors and discrepancies were resolved by discussion. For studies reported in Chinese, authors who are native in the Chinese language performed the data extraction and translated them into English for collation. The following information was extracted: (1) study characteristics: study design, study site and recruitment period; (2) patient and disease characteristics: number of patients, age, gender, primary tumour location and tumour stage; (3) study intervention: type of CIK therapy and non-CIK anti-cancer therapy received; (4) clinical efficacy endpoints: overall survival (OS), progression-free survival (PFS), 1-, 3- and 5-year OS rates, 1-, 3-, 5-year PFS rates, and overall response rate (ORR); and (6) toxicity.

For studies where patients received curative-intent treatment, disease-free survival (DFS) and DFS rates were extracted as PFS and PFS rates. Risk of bias was assessed for the following domains and graded as high, low or unclear: (1) random sequence generation; (2) allocation concealment; (3) blinding of participants and personnel; (4) blinding of outcome assessment; (5) imbalance in baseline characteristics; (6) incomplete outcome data; and (7) uniformity of non-CIK/DC-CIK anti-cancer treatment administered between intervention and control arms.

### Data synthesis and analysis

Review Manager 5.4.1^21^ was used for pooling data at the study level and statistical analysis. For the multi-intervention-arm study, the control arm was split equally into each intervention arm, so that each pair-wise comparison can be entered separately. Pooled estimates of effect were expressed as a hazard ratio (HR) calculated using an inverse variance model for OS and PFS, and risk ratios (RRs) were calculated using Mantel-Haenszel model for survival rates and ORRs. When individual studies did not describe OS and/or PFS HRs and associated 95% confidence intervals (95% CI), they were estimated from the published Kaplan-Meier curves using a previously described method^22,23^. The HR and 95% CI were estimated under the assumption for Gaussian distribution for the study that reported median PFS with a p-value^24^.

As heterogeneity due to clinical diversity was expected to be high, a random-effects model was used for all the quantitative analyses performed in this review. Heterogeneity across studies was further assessed by visual inspection and statistically using Chi-square (Chi^2^) test and I-square (I^2^) statistics for each analysis. A p-value threshold of 0.10 was employed to determine statistical significance for Chi^2^ test and I^2^ of 30% or less was considered to be a low degree of heterogeneity, 30% to 60% to be a moderate degree, and 60% or more to be a high degree.

Subgroup analyses were carried out on OS and PFS endpoints to investigate possible sources of heterogeneity. The following subgroup analyses were performed in this review: (1) quality of study design: randomised studies versus non-randomised studies; (2) cancer staging: Stage I-III after resection of primary versus Stage IV (unresectable, metastatic, or recurrent) CRC; (3) CIK therapy type: CIK therapy versus DC-CIK therapy; and (4) CIK therapy administration timing in relation to other anti-cancer therapy: concurrent vs sequential. The subgroup interactions were tested by using the formal statistical test, Chi^2^ test, with significance set at 10%.

## Results

### Search results

Through our electronic search 333 records were identified: 129 from Embase, 38 from MEDLINE, 60 from CNKI and 106 from Wanfang Data. After removing duplicate publications and studies which titles and/or abstracts indicated were ineligible, 106 records were assessed in detail. An additional 36 records were excluded for: only a single study arm; lack of information on clinical efficacy endpoints of interest; overlapping patient cohorts with another publication; being unable to extract data specific to CRC patients; inability to locate original abstracts or full-text articles; patients in all study arms receiving CIK therapy; and patients in the control arm being healthy subjects. Thus, 70 studies containing 16 English^17,19,25-38^ and 44 Chinese^24,39-91^ language articles were selected for study synthesis (Figure 1).

**Figure 1.**
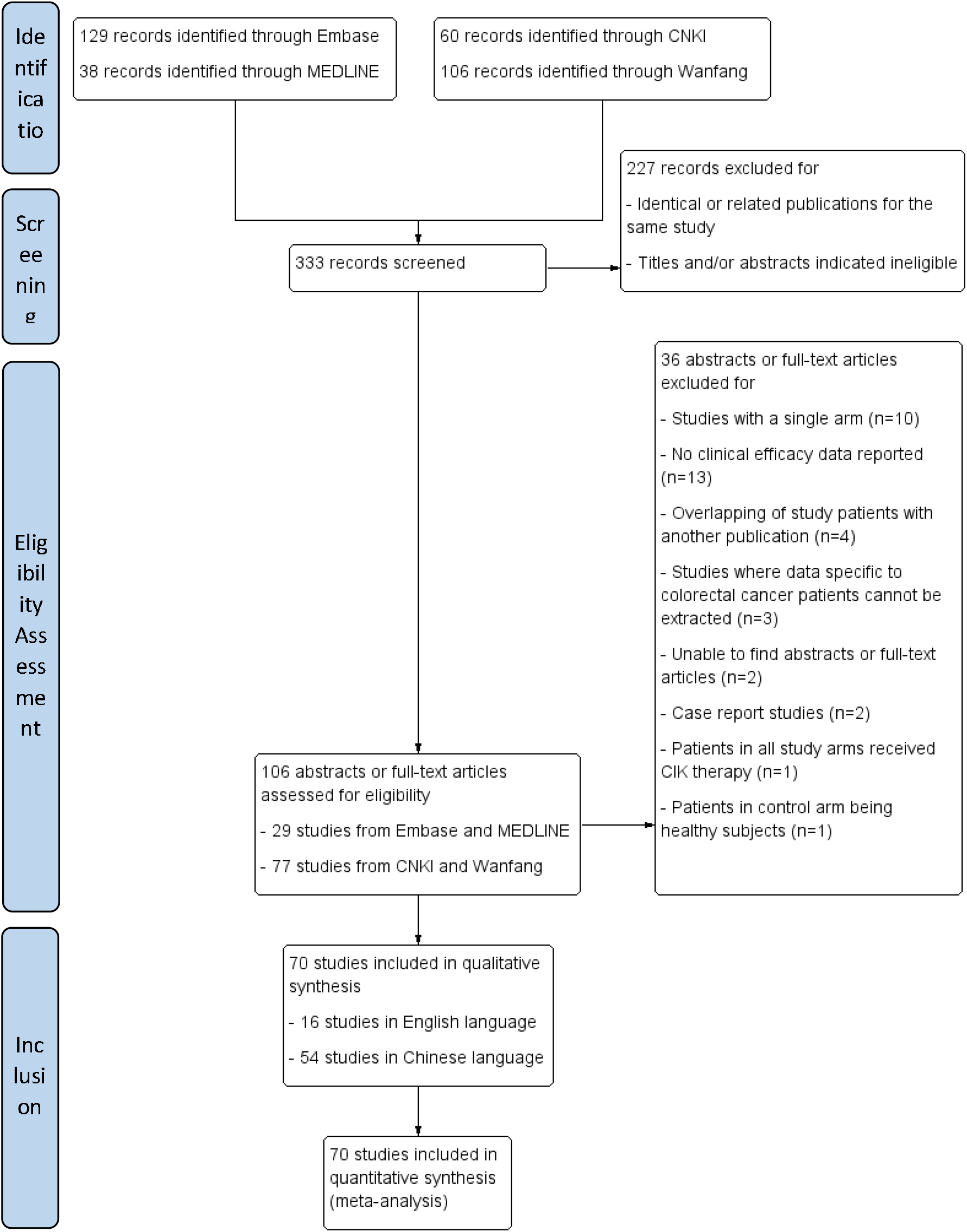
Flow chart of study selection.

### Study and patient characteristics

Standardised study cohorts are summarised in Table 1. Two studies^33,92^ were abstracts with the rest being full-text articles. All studies were single-centre studies performed in mainland China. Fifty-four studies^24-26,29,32,35,36,39-42,44-53,55,57-63,65-75,77,78,80-83,85-91^ were prospective and 15 studies^17,19,28,31,33,34,37,38,43,54,56,64,76,79,84^ were retrospective in nature. Of the prospective studies, 38^25,26,29,32,35,36,39,40,44-47,49,50,52,53,57,59,61-63,65-69,72-75,80-83,85,87,89,91^ were randomised controlled studies.

**Table 1.**
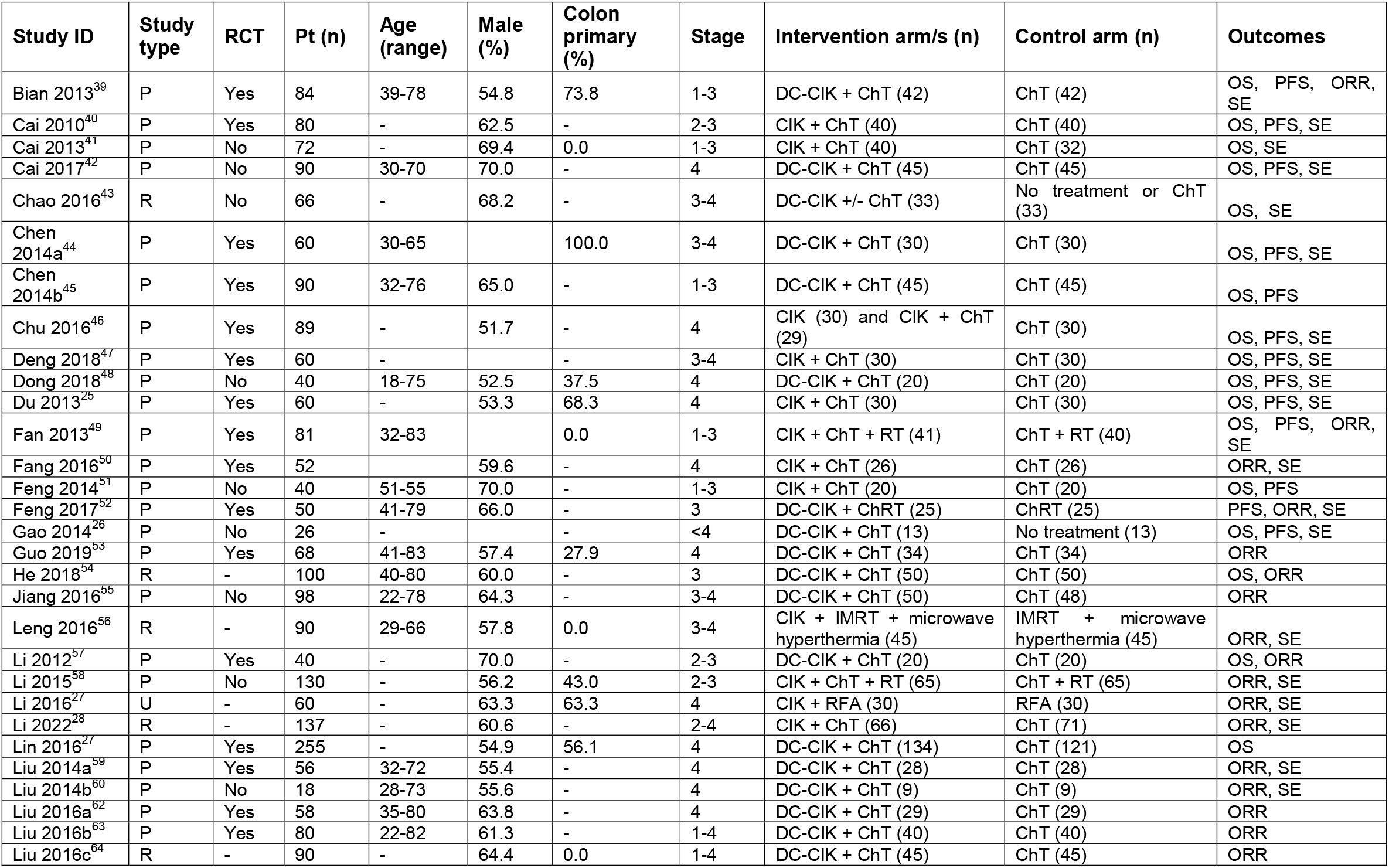
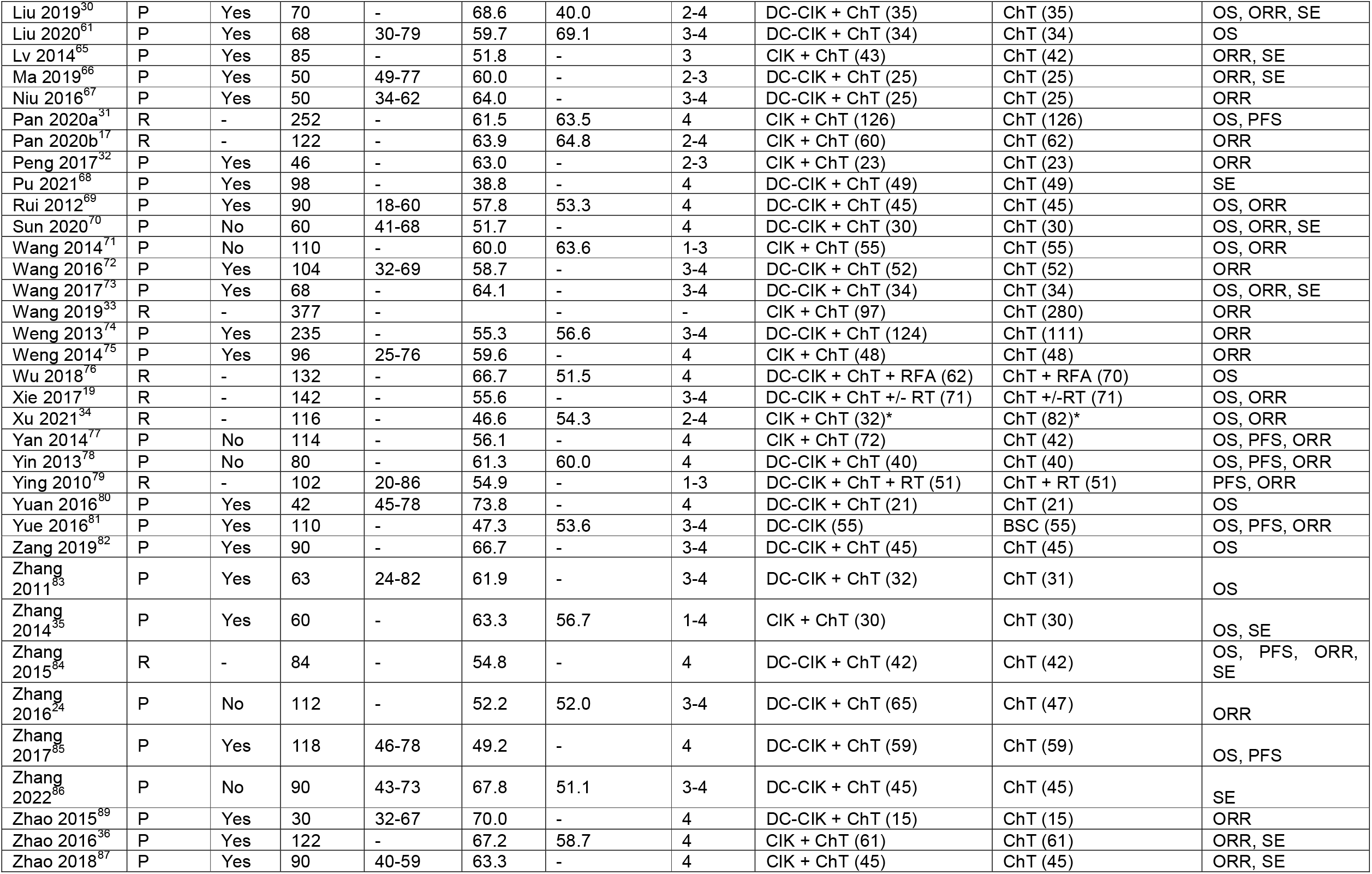
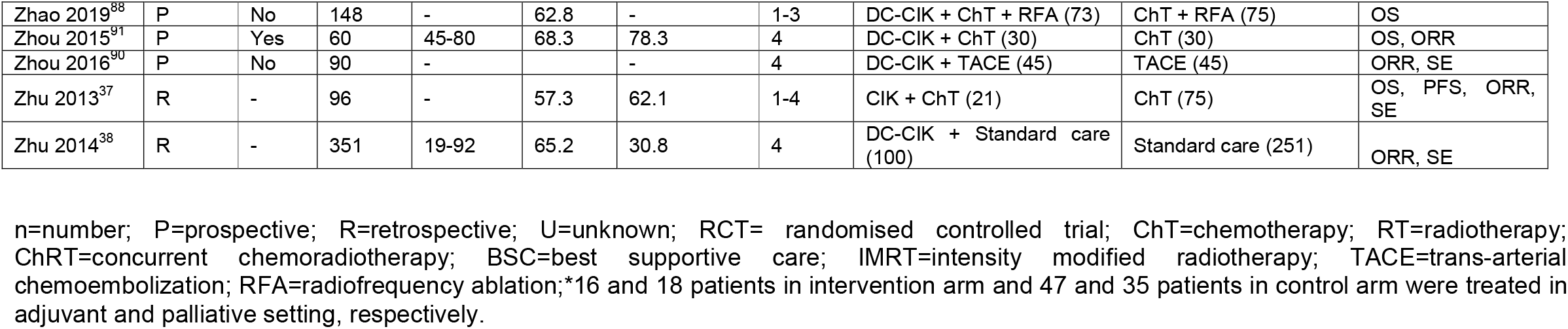
Summary of included studies

| Study ID | Study type | RCT | Pt (n) | Age (range) | Male (%) | Colon primary (%) | Stage | Intervention arm/s (n) | Control arm (n) | Outcomes |
| --- | --- | --- | --- | --- | --- | --- | --- | --- | --- | --- |
| Bian 2013<sup>39</sup> | P | Yes | 84 | 39-78 | 54.8 | 73.8 | 1-3 | DC-CIK + ChT (42) | ChT (42) | OS, PFS, ORR, SE |
| Cai 2010<sup>40</sup> | P | Yes | 80 | - | 62.5 | - | 2-3 | CIK + ChT (40) | ChT (40) | OS, PFS, SE |
| Cai 2013<sup>41</sup> | P | No | 72 | - | 69.4 | 0.0 | 1-3 | CIK + ChT (40) | ChT (32) | OS, SE |
| Cai 2017<sup>42</sup> | P | No | 90 | 30-70 | 70.0 | - | 4 | DC-CIK + ChT (45) | ChT (45) | OS, PFS, SE |
| Chao 2016<sup>43</sup> | R | No | 66 | - | 68.2 | - | 3-4 | DC-CIK +/- ChT (33) | No treatment or ChT (33) | OS, SE |
| Chen 2014a<sup>44</sup> | P | Yes | 60 | 30-65 |  | 100.0 | 3-4 | DC-CIK + ChT (30) | ChT (30) | OS, PFS, SE |
| Chen 2014b<sup>45</sup> | P | Yes | 90 | 32-76 | 65.0 | - | 1-3 | DC-CIK + ChT (45) | ChT (45) | OS, PFS |
| Chu 2016<sup>46</sup> | P | Yes | 89 | - | 51.7 | - | 4 | CIK (30) and CIK + ChT (29) | ChT (30) | OS, PFS, SE |
| Deng 2018<sup>47</sup> | P | Yes | 60 | - |  | - | 3-4 | CIK + ChT (30) | ChT (30) | OS, PFS, SE |
| Dong 2018<sup>48</sup> | P | No | 40 | 18-75 | 52.5 | 37.5 | 4 | DC-CIK + ChT (20) | ChT (20) | OS, PFS, SE |
| Du 2013<sup>25</sup> | P | Yes | 60 | - | 53.3 | 68.3 | 4 | CIK + ChT (30) | ChT (30) | OS, PFS, SE |
| Fan 2013<sup>49</sup> | P | Yes | 81 | 32-83 |  | 0.0 | 1-3 | CIK + ChT + RT (41) | ChT + RT (40) | OS, PFS, ORR, SE |
| Fang 2016<sup>50</sup> | P | Yes | 52 |  | 59.6 | - | 4 | CIK + ChT (26) | ChT (26) | ORR, SE |
| Feng 2014<sup>51</sup> | P | No | 40 | 51-55 | 70.0 | - | 1-3 | CIK + ChT (20) | ChT (20) | OS, PFS |
| Feng 2017<sup>52</sup> | P | Yes | 50 | 41-79 | 66.0 | - | 3 | DC-CIK + ChRT (25) | ChRT (25) | PFS, ORR, SE |
| Gao 2014<sup>26</sup> | P | No | 26 | - |  | - | <4 | DC-CIK + ChT (13) | No treatment (13) | OS, PFS, SE |
| Guo 2019<sup>53</sup> | P | Yes | 68 | 41-83 | 57.4 | 27.9 | 4 | DC-CIK + ChT (34) | ChT (34) | ORR |
| He 2018<sup>54</sup> | R | - | 100 | 40-80 | 60.0 | - | 3 | DC-CIK + ChT (50) | ChT (50) | OS, ORR |
| Jiang 2016<sup>55</sup> | P | No | 98 | 22-78 | 64.3 | - | 3-4 | DC-CIK + ChT (50) | ChT (48) | ORR |
| Leng 2016<sup>56</sup> | R | - | 90 | 29-66 | 57.8 | 0.0 | 3-4 | CIK + IMRT + microwave hyperthermia (45) | IMRT + microwave hyperthermia (45) | ORR, SE |
| Li 2012<sup>57</sup> | P | Yes | 40 | - | 70.0 | - | 2-3 | DC-CIK + ChT (20) | ChT (20) | OS, ORR |
| Li 2015<sup>58</sup> | P | No | 130 | - | 56.2 | 43.0 | 2-3 | CIK + ChT + RT (65) | ChT + RT (65) | ORR, SE |
| Li 2016<sup>27</sup> | U | - | 60 | - | 63.3 | 63.3 | 4 | CIK + RFA (30) | RFA (30) | ORR, SE |
| Li 2022<sup>28</sup> | R | - | 137 | - | 60.6 | - | 2-4 | CIK + ChT (66) | ChT (71) | ORR, SE |
| Lin 2016<sup>27</sup> | P | Yes | 255 | - | 54.9 | 56.1 | 4 | DC-CIK + ChT (134) | ChT (121) | OS |
| Liu 2014a<sup>59</sup> | P | Yes | 56 | 32-72 | 55.4 | - | 4 | DC-CIK + ChT (28) | ChT (28) | ORR, SE |
| Liu 2014b<sup>60</sup> | P | No | 18 | 28-73 | 55.6 | - | 4 | DC-CIK + ChT (9) | ChT (9) | ORR, SE |
| Liu 2016a<sup>62</sup> | P | Yes | 58 | 35-80 | 63.8 | - | 4 | DC-CIK + ChT (29) | ChT (29) | ORR |
| Liu 2016b<sup>63</sup> | P | Yes | 80 | 22-82 | 61.3 | - | 1-4 | DC-CIK + ChT (40) | ChT (40) | ORR |
| Liu 2016c<sup>64</sup> | R | - | 90 | - | 64.4 | 0.0 | 1-4 | DC-CIK + ChT (45) | ChT (45) | ORR |
| Liu 2019<sup>30</sup> | P | Yes | 70 | - | 68.6 | 40.0 | 2-4 | DC-CIK + ChT (35) | ChT (35) | OS, ORR, SE |
| Liu 2020<sup>61</sup> | P | Yes | 68 | 30-79 | 59.7 | 69.1 | 3-4 | DC-CIK + ChT (34) | ChT (34) | OS |
| Lv 2014<sup>65</sup> | P | Yes | 85 | - | 51.8 | - | 3 | CIK + ChT (43) | ChT (42) | ORR, SE |
| Ma 2019<sup>66</sup> | P | Yes | 50 | 49-77 | 60.0 | - | 2-3 | DC-CIK + ChT (25) | ChT (25) | ORR, SE |
| Niu 2016<sup>67</sup> | P | Yes | 50 | 34-62 | 64.0 | - | 3-4 | DC-CIK + ChT (25) | ChT (25) | ORR |
| Pan 2020a<sup>31</sup> | R | - | 252 | - | 61.5 | 63.5 | 4 | CIK + ChT (126) | ChT (126) | OS, PFS |
| Pan 2020b<sup>17</sup> | R | - | 122 | - | 63.9 | 64.8 | 2-4 | CIK + ChT (60) | ChT (62) | ORR |
| Peng 2017<sup>32</sup> | P | Yes | 46 | - | 63.0 | - | 2-3 | CIK + ChT (23) | ChT (23) | ORR |
| Pu 2021<sup>68</sup> | P | Yes | 98 | - | 38.8 | - | 4 | DC-CIK + ChT (49) | ChT (49) | SE |
| Rui 2012<sup>69</sup> | P | Yes | 90 | 18-60 | 57.8 | 53.3 | 4 | DC-CIK + ChT (45) | ChT (45) | OS, ORR |
| Sun 2020<sup>70</sup> | P | No | 60 | 41-68 | 51.7 | - | 4 | DC-CIK + ChT (30) | ChT (30) | OS, ORR, SE |
| Wang 2014<sup>71</sup> | P | No | 110 | - | 60.0 | 63.6 | 1-3 | CIK + ChT (55) | ChT (55) | OS, ORR |
| Wang 2016<sup>72</sup> | P | Yes | 104 | 32-69 | 58.7 | - | 3-4 | DC-CIK + ChT (52) | ChT (52) | ORR |
| Wang 2017<sup>73</sup> | P | Yes | 68 | - | 64.1 | - | 3-4 | DC-CIK + ChT (34) | ChT (34) | OS, ORR, SE |
| Wang 2019<sup>33</sup> | R | - | 377 | - | - | - | - | CIK + ChT (97) | ChT (280) | ORR |
| Weng 2013<sup>74</sup> | P | Yes | 235 | - | 55.3 | 56.6 | 3-4 | DC-CIK + ChT (124) | ChT (111) | ORR |
| Weng 2014<sup>75</sup> | P | Yes | 96 | 25-76 | 59.6 | - | 4 | CIK + ChT (48) | ChT (48) | ORR |
| Wu 2018<sup>76</sup> | R | - | 132 | - | 66.7 | 51.5 | 4 | DC-CIK + ChT + RFA (62) | ChT + RFA (70) | OS |
| Xie 2017<sup>19</sup> | R | - | 142 | - | 55.6 | - | 3-4 | DC-CIK + ChT +/- RT (71) | ChT +/-RT (71) | OS, ORR |
| Xu 2021<sup>34</sup> | R | - | 116 | - | 46.6 | 54.3 | 2-4 | CIK + ChT (32)* | ChT (82)* | OS, ORR |
| Yan 2014<sup>77</sup> | P | No | 114 | - | 56.1 | - | 4 | CIK + ChT (72) | ChT (42) | OS, PFS, ORR |
| Yin 2013<sup>78</sup> | P | No | 80 | - | 61.3 | 60.0 | 4 | DC-CIK + ChT (40) | ChT (40) | OS, PFS, ORR |
| Ying 2010<sup>79</sup> | R | - | 102 | 20-86 | 54.9 | - | 1-3 | DC-CIK + ChT + RT (51) | ChT + RT (51) | PFS, ORR |
| Yuan 2016<sup>80</sup> | P | Yes | 42 | 45-78 | 73.8 | - | 4 | DC-CIK + ChT (21) | ChT (21) | OS |
| Yue 2016<sup>81</sup> | P | Yes | 110 | - | 47.3 | 53.6 | 3-4 | DC-CIK (55) | BSC (55) | OS, PFS, ORR |
| Zang 2019<sup>82</sup> | P | Yes | 90 | - | 66.7 | - | 3-4 | DC-CIK + ChT (45) | ChT (45) | OS |
| Zhang 2011<sup>83</sup> | P | Yes | 63 | 24-82 | 61.9 | - | 3-4 | DC-CIK + ChT (32) | ChT (31) | OS |
| Zhang 2014<sup>35</sup> | P | Yes | 60 | - | 63.3 | 56.7 | 1-4 | CIK + ChT (30) | ChT (30) | OS, SE |
| Zhang 2015<sup>84</sup> | R | - | 84 | - | 54.8 | - | 4 | DC-CIK + ChT (42) | ChT (42) | OS, PFS, ORR, SE |
| Zhang 2016<sup>24</sup> | P | No | 112 | - | 52.2 | 52.0 | 3-4 | DC-CIK + ChT (65) | ChT (47) | ORR |
| Zhang 2017<sup>85</sup> | P | Yes | 118 | 46-78 | 49.2 | - | 4 | DC-CIK + ChT (59) | ChT (59) | OS, PFS |
| Zhang 2022<sup>86</sup> | P | No | 90 | 43-73 | 67.8 | 51.1 | 3-4 | DC-CIK + ChT (45) | ChT (45) | SE |
| Zhao 2015<sup>89</sup> | P | Yes | 30 | 32-67 | 70.0 | - | 4 | DC-CIK + ChT (15) | ChT (15) | ORR |
| Zhao 2016<sup>36</sup> | P | Yes | 122 | - | 67.2 | 58.7 | 4 | CIK + ChT (61) | ChT (61) | ORR, SE |
| Zhao 2018<sup>87</sup> | P | Yes | 90 | 40-59 | 63.3 | - | 4 | CIK + ChT (45) | ChT (45) | ORR, SE |
| Zhao 2019<sup>88</sup> | P | No | 148 | - | 62.8 | - | 1-3 | DC-CIK + ChT + RFA (73) | ChT + RFA (75) | OS |
| Zhou 2015<sup>91</sup> | P | Yes | 60 | 45-80 | 68.3 | 78.3 | 4 | DC-CIK + ChT (30) | ChT (30) | OS, ORR |
| Zhou 2016<sup>90</sup> | P | No | 90 | - | - | - | 4 | DC-CIK + TACE (45) | TACE (45) | ORR, SE |
| Zhu 2013<sup>37</sup> | R | - | 96 | - | 57.3 | 62.1 | 1-4 | CIK + ChT (21) | ChT (75) | OS, PFS, ORR, SE |
| Zhu 2014<sup>38</sup> | R | - | 351 | 19-92 | 65.2 | 30.8 | 4 | DC-CIK + Standard care (100) | Standard care (251) | ORR, SE |

n=number; P=prospective; R=retrospective; U=unknown; RCT= randomised controlled trial; ChT=chemotherapy; RT=radiotherapy; ChRT=concurrent chemoradiotherapy; BSC=best supportive care; IMRT=intensity modified radiotherapy; TACE=trans-arterial chemoembolization; RFA=radiofrequency ablation;\*16 and 18 patients in intervention arm and 47 and 35 patients in control arm were treated in adjuvant and palliative setting, respectively.

Overall, 6,743 CRC patients, 3,203 in CIK therapy (intervention) arm and 3,540 in non-CIK therapy (control) arm were available for analysis. The median age ranged from 43.2 to 80.0 years old with the youngest being 18 and the oldest being 92 years old. For studies which provided the patient’s gender, 3,592 out of 6,017 patients (59.7%) were males. Primary tumour location was reported in thirty studies, with 1,657 colon and 1,744 rectum cancer patients. CRC patients diagnosed with all cancer stages were considered for analysis. Three studies evaluated purely Stage III CRC patients^52,54,65^, while 29 studies evaluated Stage IV CRC patients^25,27,29,31,36,38,42,46,48,50,53,59,60,62,68-70,75-78,80,84,85,87,89-91^. The remaining studies considered patients with multiple stages. Among patients with known cancer stages, 3109 (66.6%) of them had Stage IV disease, comprising the largest group followed by 1,148 patients (24.5%) with Stage III disease, 375 patients with Stage II disease and 46 patients with Stage I disease. Cancer staging for the remaining 1,672 patients were either unknown or reported in ranges.

### Interventions

In 25 studies^17,25,27,28,31-37,40,41,46,47,49-51,56,58,65,71,75,77,87^, patients in the intervention arm received CIK therapy, while in 45 studies^19,24,26,29,30,38,39,42-45,48,5 2-55,57,59-64,66-70,72-74,76,78-86,88-91^ DC-CIK therapy was administered. Chemotherapy was the most common co-treatment with CIK or DC-CIK therapy, being utilised in 66 studies^17,19,24-26,28-55,57-80,82-90^. The most commonly utilised chemotherapy regimens were FOLFOX and XELOX, being administered in 43^17,24,28,29,31,32,34-37,39,41,43,45,48,50,51,55,57-66,69-71,73-76,78,79,82-85,88,91^ and 24^17,25,26,29,31,34,35,37,40-42,47,58,67,70,72,86,87,89^ studies, respectively.

Other less commonly used regimens included 5-fluorouracil monotherapy in 6 studies^29,34,37,41,55,79^, capecitabine monotherapy in 7 studies^17,34,37,55,58,77,84^, FOLFIRI in 8 studies^19,31,43,48,76,78,83,88^ and FOLFOXIRI in 2 studies^76,88^. In total, 2,847 patients in the intervention arm and 3,033 patients in the control arm were confirmed to have received chemotherapy as a part of study intervention. In 10 studies, local therapy was administered together with CIK/DC-CIK therapy: radiofrequency ablation in 3 studies^27,76,88^, radiotherapy in 6 studies^19,49,52,56,58,79^, transarterial chemoembolization (TACE) in 1 study^90^ and microwave hyperthermia in one study^56^. In 2 studies, some or all patients in the intervention arm received CIK/DC-CIK therapy alone^43,81^.

### Risk of bias assessment

Risk of bias assessment is shown in Supplementary Figure 1. Among the 38 studies reported to be prospective randomised controlled studies, only 9 studies^26,32,36,45,61,68,72,80,91^ described the method of randomisation and no study discussed allocation concealment. None of the included studies provided clarity on blinding of patients, study personnel or investigators. However, it was considered unlikely that a lack of blinding would affect the clinical efficacy endpoints evaluated in the review, namely OS, PFS, OS rate and PFS rate, and ORR. All the studies were thus assessed to be at low risk of performance and detection bias secondary to insufficient blinding. Demographic and clinical characteristics of patients were generally well-balanced across the studies. Four studies^26,33,51,60^ were at unclear risk of selection bias due to a lack of patient characteristics information across treatment arms. Imbalance in age, cancer stage and history of primary cancer resection was noted for 3 studies^17,37,38^ and they were similarly assessed to be at unclear risk of selection bias. Unclear risk of performance bias due to uncertainty around uniformity of non-CIK/DC-CIK treatment across the intervention and control arms was identified in 21 studies^26,29,31,33,34,38,43,44,48,52,53,55,58,62,76,79-81,83,84,88^ with all the studies except one failing to adequately describe study interventions or the proportion of patients receiving various interventions. In the remaining 1 study^81^, patients in the intervention arm received DC-CIK therapy alone, while those in the control arm received best supportive care. The risk of attrition bias was rated unclear for 18 studies^17,19,25,27,29,31-33,35-38,40,42,68,79,83,85,90^ which did not reveal the number of patients lost in follow-up and for 1 study^37^ in which 18.8% of patients withdrew from the study prematurely.

### Overall survival and progression-free survival

There were 26 studies^17,19,26,28,31,32,34,37,40,41,43,45,49,50,55,57-59,62,65,69,77,79,81,83,88^ involving 3,303 patients which contributed data to the meta-analysis on OS (Figure 2A). The pooled HR was 0.59 (95% CI 0.53-0.65) indicating OS benefit of CIK/DC-CIK therapy over the control arm. Heterogeneity among the studies was low (I^2^=11%, p=0.30). For PFS, 20 studies^19,25-29,31,33-37,58,64,74,76,78,79,84,86^ involving 2,593 patients contributed the data to the meta-analysis (Figure 2B). The pooled HR was 0.55 (95%CI 0.47-0.63), again favouring CIK/DC-CIK therapy. Heterogeneity among the studies was moderate (I^2^=54%, p=0.002).

**Figure 2.**
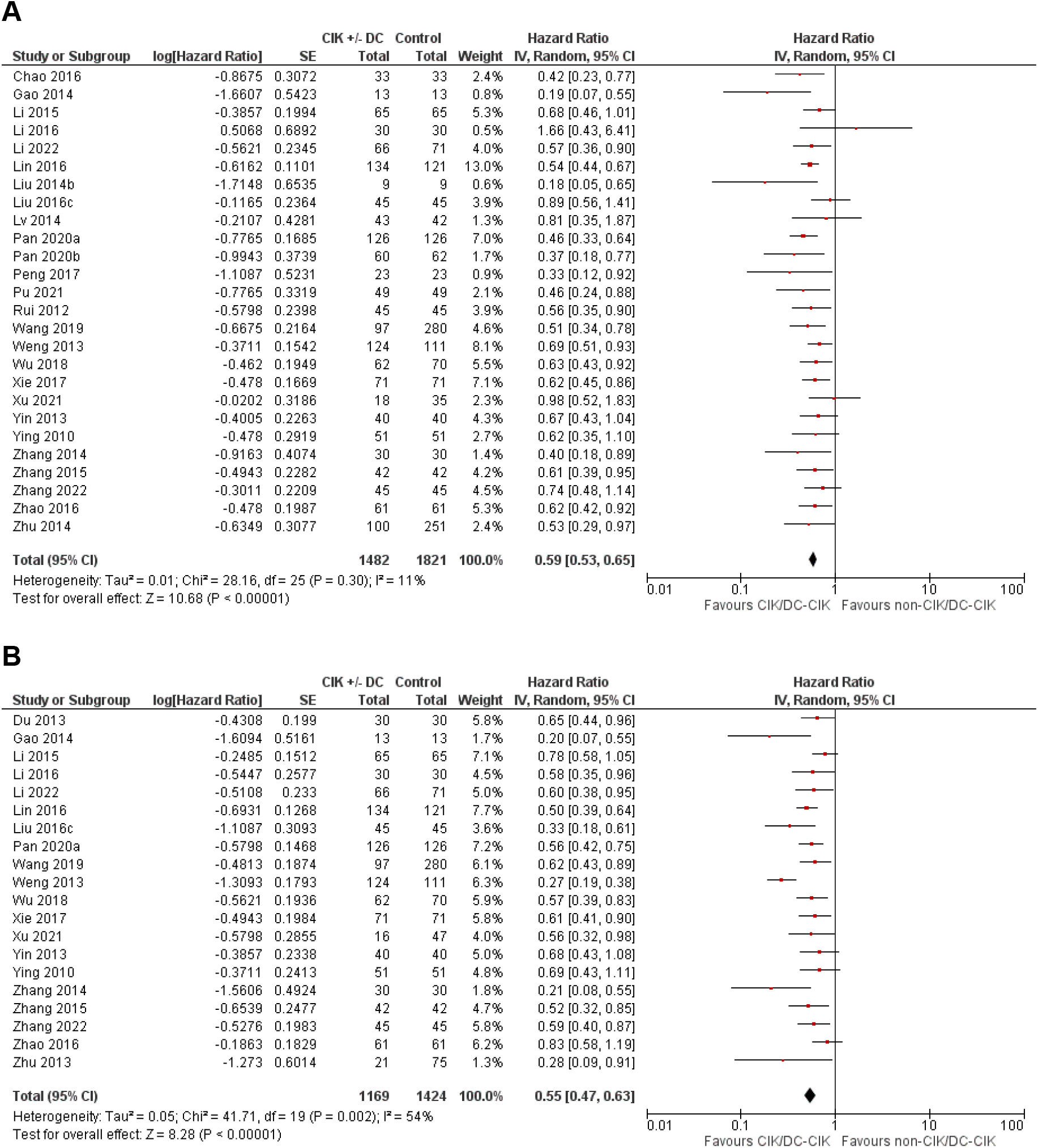
Comparison of CIK/DC-CIK therapy versus non-CIK/DC-CIK therapy for (A) overall survival (OS) and (B) progression-free survival (PFS). Twenty-six studies involving 3,303 patients and twenty studies involving 2,593 patients contributed data to OS and PFS analysis respectively. CIK, cytokine-induced killer cell; DC, dendritic cell.

### Overall survival rates

In total, 27 (2,459 patients)^17,19,26,28,31,32,34,37,40,41,43,45,49-51,55,57-59,62,65,69,77,79,81,83,88^, 19 (2,167 patients)^17,19,26-28,31,33,34,36,37,40,41,43,50,51,58,69,79,88^ and 10 (1,401 patients) ^17,19,26,28,31,33,34,37,43,58^ studies contributed data for 1-year, 3-year and 5-year OS rate meta-analyses respectively (Supplementary Figure 2). The pooled RR for all the analyses favoured CIK/DC-CIK therapy. The 1-year OS rate was 91.7% in the intervention arm and 79.4% in the control arm with a pooled RR of 0.47 (95% CI 0.32-0.67). Heterogeneity among the studies was moderate (I^2^=51%, p=0.002). The 3-year OS rate was 67.7% in the intervention arm and 51.8% in the control arm with the pooled RR of 0.67 (95% CI 0.59-0.77). There was a moderate level of heterogeneity among the studies (I^2^ = 32%, p=0.09). The 5-year OS rate was 61.2% in the intervention arm and 45.5% in the control arm with RR of 0.69 (95% CI 0.54-0.88). Heterogeneity among the studies was high (I^2^ = 73%, P = 0.0001).

### Progression-free survival rates

We identified 10 (1,166 patients)^17,19,26,28,31,34,37,38,58,79^, 10 (1,156 patients)^17,19,26-28,31,34,36,58,79^ and 7 (872 patients) studies^17,19,26,28,31,34,58^ that contributed data for meta-analysis on 1-year, 3-year and 5-year PFS rates respectively (Supplementary Figure 3). All the analyses indicated the superiority of CIK/DC-CIK therapy over non-CIK/DC-CIK therapy. The observed 1-year PFS rate was 86.5% in the intervention arm and 68.1% in the control with the pooled RR of 0.43 (95% CI 0.33-0.55). Heterogeneity among the studies was low (I^2^ = 0%, p=0.48). The 3-year PFS rate was 47.8% in the intervention arm and 30.5% in the control arm. The pooled RR was 0.76 (95% CI 0.66-0.87) and heterogeneity among the studies was moderate (I^2^=53%, p=0.02). At 5 years, PFS rate was 46.0% in the intervention arm and 25.9% in the control arm. The pooled RR was 0.71 (95% CI 0.59-0.87) and heterogeneity among the studies was high (I^2^=68%, p=0.005).

### Overall response rate

The ORR was 58.7% in the intervention (CIK/DC-CIK) and 39.8% in the control (non CIK/DC-CIK) arm for 3,860 patients from 45 studies^24,25,30,36,39,42,44-49,52-56,61-68,70-78,80-85,87-91^ (Figure 3). The pooled RR was 0.65 (95% CI 0.57-0.74) and heterogeneity among the studies was high (I^2^ =85%, p<0.00001).

**Figure 3.**
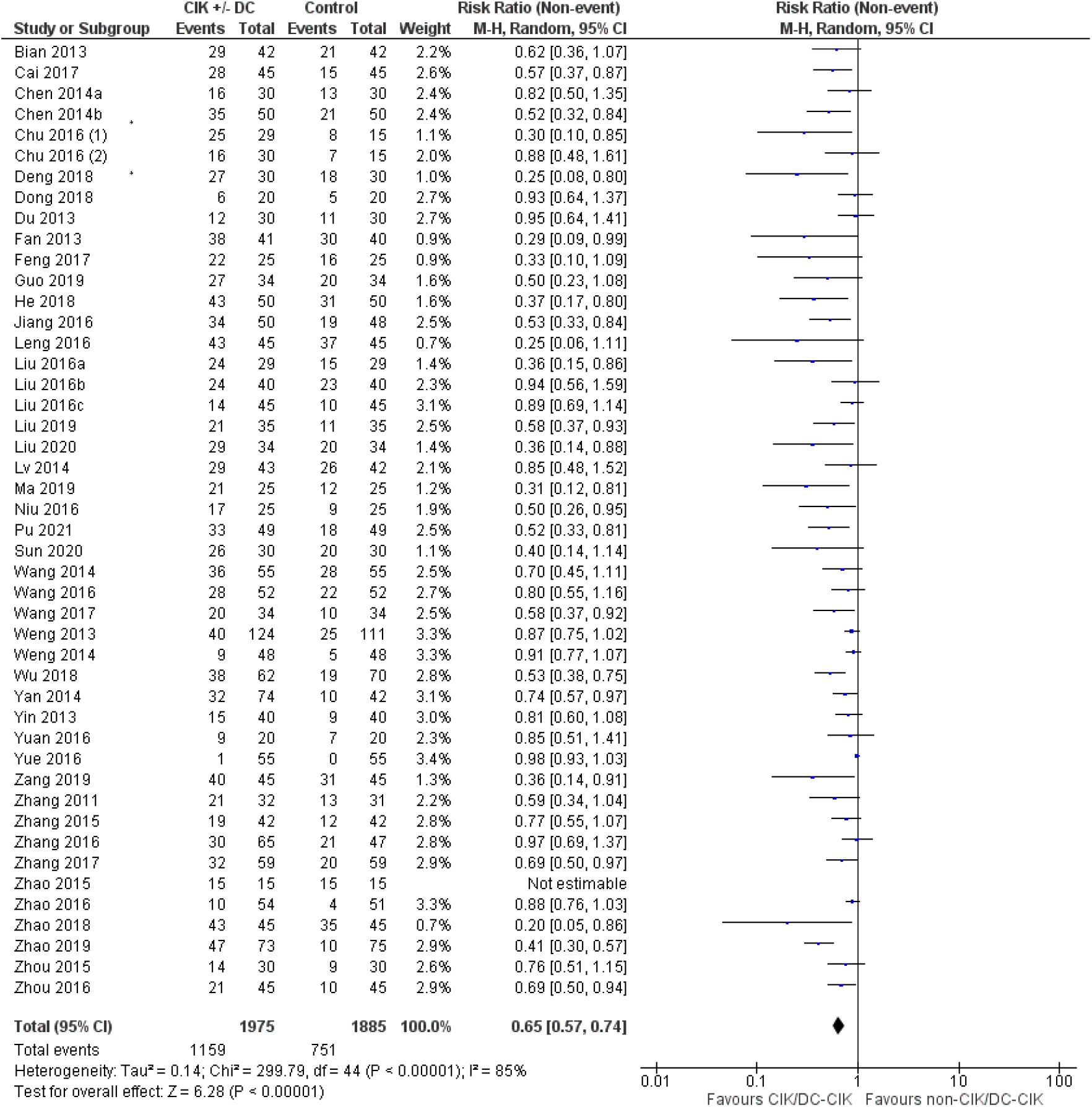
CIK/DC-CIK therapy versus non-CIK/DC-CIK therapy for overall response rate (ORR). Forty-five studies involving 3,860 patients contributed data to ORR analysis. *Study Chu 2016 appears twice in the figure as it contained 3 treatment arms and data were entered separately for CIK + chemotherapy versus chemotherapy (Chu 2016 (1)) and CIK versus chemotherapy (Chu 2016 (2)) by splitting the chemotherapy group into 2 subgroups, one for each CIK + chemotherapy and CIK treatment. CIK, cytokine-induced killer cell; DC, dendritic cell.

### Toxicity

Toxicity during the study intervention was reported by 31 studies with the majority of the data being provided in a descriptive manner. Two studies^33,87^ compared the rate of any adverse events between the treatment arms and 11 studies^29,36,39,41,42,44,56,61,62,70,72,89^ reported adverse events of interest for each arm. Many of the described side effects were thought to be related to chemotherapy administered together with CIK/DC-CIK therapy, including bone marrow suppression, nausea, vomiting, neuropathy, diarrhoea and liver dysfunction. Meta-analysis undertaken indicated equivalent adverse event rate from CIK/DC-CIK and non-CIK/DC-CIK therapy (HR=0.59, 95% CI 016-2.25) with the pooled adverse event rate of 53.5% and 68.3%, respectively (Supplementary Figure 4). Heterogeneity was high between the studies (I^2^=80%, p=0.02). Fever was the most frequently reported adverse event associated with CIK/DC-CIK infusion, affecting 6.7% to 29.9% of patients receiving CIK/DC-CIK therapy. Fever, in general, spontaneously resolved or only required symptomatic management.

### Subgroup analyses

Potential sources of heterogeneity were explored by performing subgroup analysis on OS and PFS by study design (randomised versus non-randomised study design), disease stage (Stage I-III versus Stage IV), CIK therapy type (CIK versus DC-CIK therapy), or timing of CIK/DC-CIK therapy administration (concurrent versus sequential with co-administered anti-cancer therapy).

#### Randomised studies versus non-randomised studies

Of the 25 studies which provided OS HRs, 8 studies^29,32,35,36,65,68,69,74^ involving 991 patients were prospective randomised studies and 17 studies^17,19,26,28,31,33,34,38,43,58,60,64,76,78,79,84,86^ involving 2,252 patients were either prospective non-randomised or retrospective studies. An OS benefit of CIK/DC-CIK therapy was demonstrated for both randomised studies (HR=0.57; 95% CI 0.50-0.66) and non-randomised studies (HR=0.59, 95% CI 0.51-0.67) (Figure 4A). A test for subgroup difference did not reach statistical significance (I^2^=0%, p=0.80). For PFS subgroup analysis, 732 patients from 5 randomised studies^25,29,35,36,74^ and 1,801 patients from 14 non-randomised studies^19,26,28,31,33,34,37,58,64,76,78,79,84,86^ were analysed. A benefit from CIK/DC-CIK therapy was again shown for both prospective randomised (HR=0.47, 95% CI 0.31-0.72) and non-randomised studies (HR=0.59, 95% CI 0.47-0.63) (Figure 4B). A test for subgroup differences was not statistically significant (I^2^=4.5%, p=0.31).

**Figure 4.**
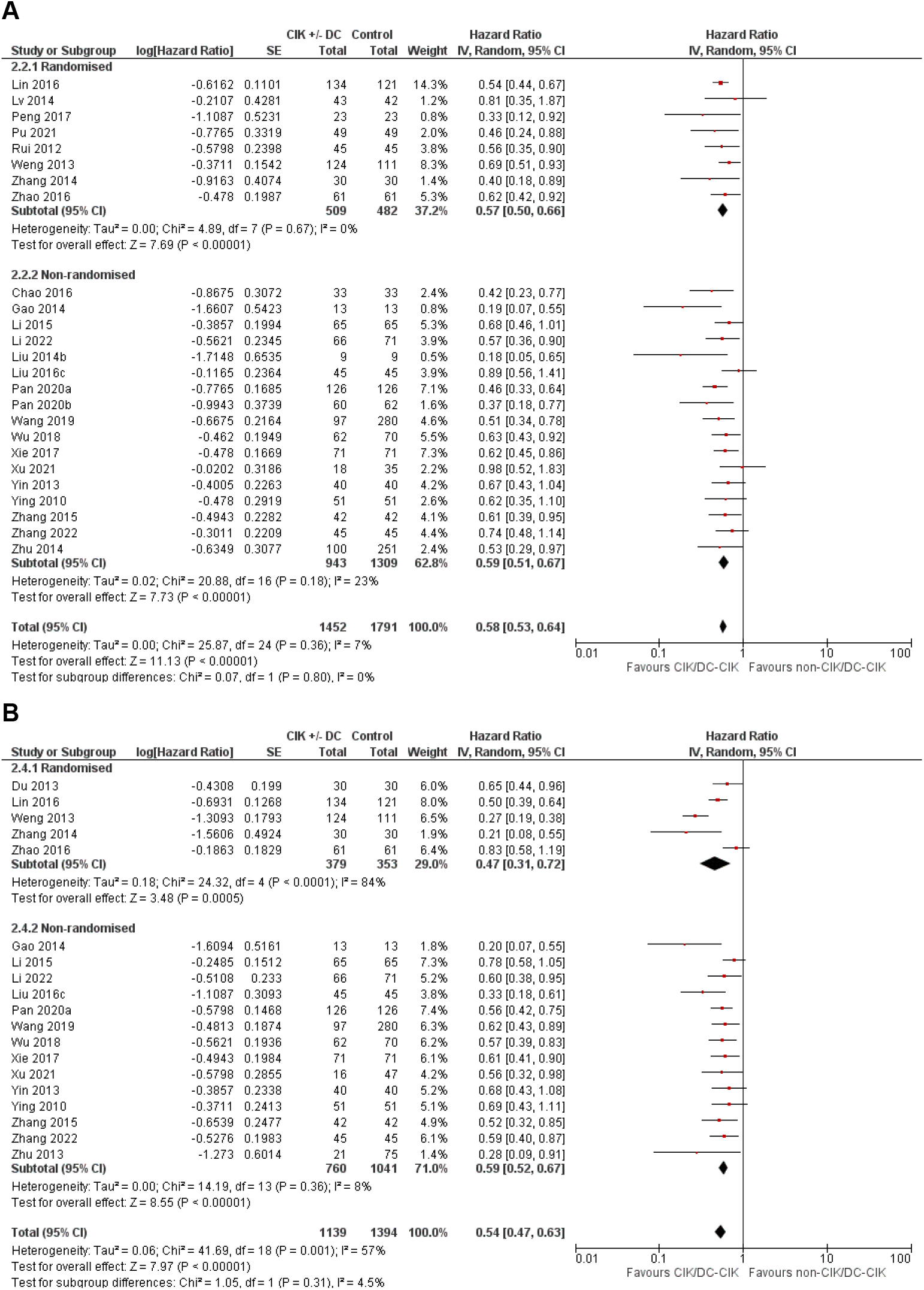
Subgroup analysis by study design for (A) overall survival (OS) and (B) progression-free survival (PFS). Twenty-five studies involving 3,243 patients and nineteen studies involving 2,533 patients contributed data to OS and PFS analysis respectively.

#### Stage I-III versus Stage IV

Four studies^32,58,65,79^ involving 363 patients with Stage I-III CRC and 12 studies^27,29,31,34,36,38,60,68,69,76,78,84^ involving 1,595 Stage IV patients contributed data to the subgroup analysis on OS by the disease stage. HR for Stage I-III patients was 0.64 (95% CI 0.48-0.85), while that for Stage IV patients was 0.57 (95% CI 0.50-0.65) and the benefit of CIK/DC-CIK therapy was observed across all stages of CRC (Supplementary Figure 5A). Test for subgroup differences failed to reach statistical significance (I^2^=0%, p=0.48), although the observed 95% CI was much narrower for Stage IV patients. For the subgroup analysis on PFS, 4 studies^26,34,58,79^ involving 321 patients with Stage I-III disease and 8 studies^25,27,29,31,36,76,78,84^ involving 1,045 Stage IV patients were analysed. A benefit from CIK/DC-CIK therapy was demonstrated for both Stage I-III (HR=0.60, 95% CI 0.40-0.88) and Stage IV disease (HR=0.59, 95% CI 0.52-0.67) (Supplementary Figure 5B). A test for subgroup difference was not statistically significant (I^2^=0%, p=0.94).

#### CIK therapy versus DC-CIK therapy

Ten studies^17,27,28,31-33,35,36,58,65^ (1,391 patients) and 16 studies^19,26,29,34,38,43,60,64,68,69,74,76,78,79,84,86^ (1,912 patients) which evaluated CIK and DC-CIK therapy respectively were assessed in the subgroup analysis on OS by type of CIK therapy. HR for studies examining CIK therapy was 0.57 (95% CI 0.47-0.69), while that for studies examining DC-CIK therapy was 0.61 (95% CI 0.54-0.69) (Figure 5A). Both types of CIK therapy were found to benefit OS. A test for subgroup differences did not reach statistical significance (I^2^=0%, p=0.58). Subgroup analysis on PFS by CIK therapy type contained 9 studies^25,27,28,31,33,35-37,58^ involving 1,294 patients, where the intervention arm contained CIK therapy, and 11 studies^19,26,29,34,64,74,76,78,79,84,86^ involving 1,299 patients, where the intervention arm contained DC-CIK therapy. PFS benefit was demonstrated for both CIK-examining (HR=0.63, 95% CI 0.53-0.74) and DC-CIK-examining studies (HR=0.50, 95% CIK 0.41-0.61) (Figure 5B). A test for subgroup differences met statistical significance (I^2^=66.5%, p=0.08) with improved HR seen for DC-CIK, although HRs for the 2 subgroups overlapped each other, suggesting that the advantage of DC-CIK over CIK therapy alone may not be clinically meaningful.

**Figure 5.**
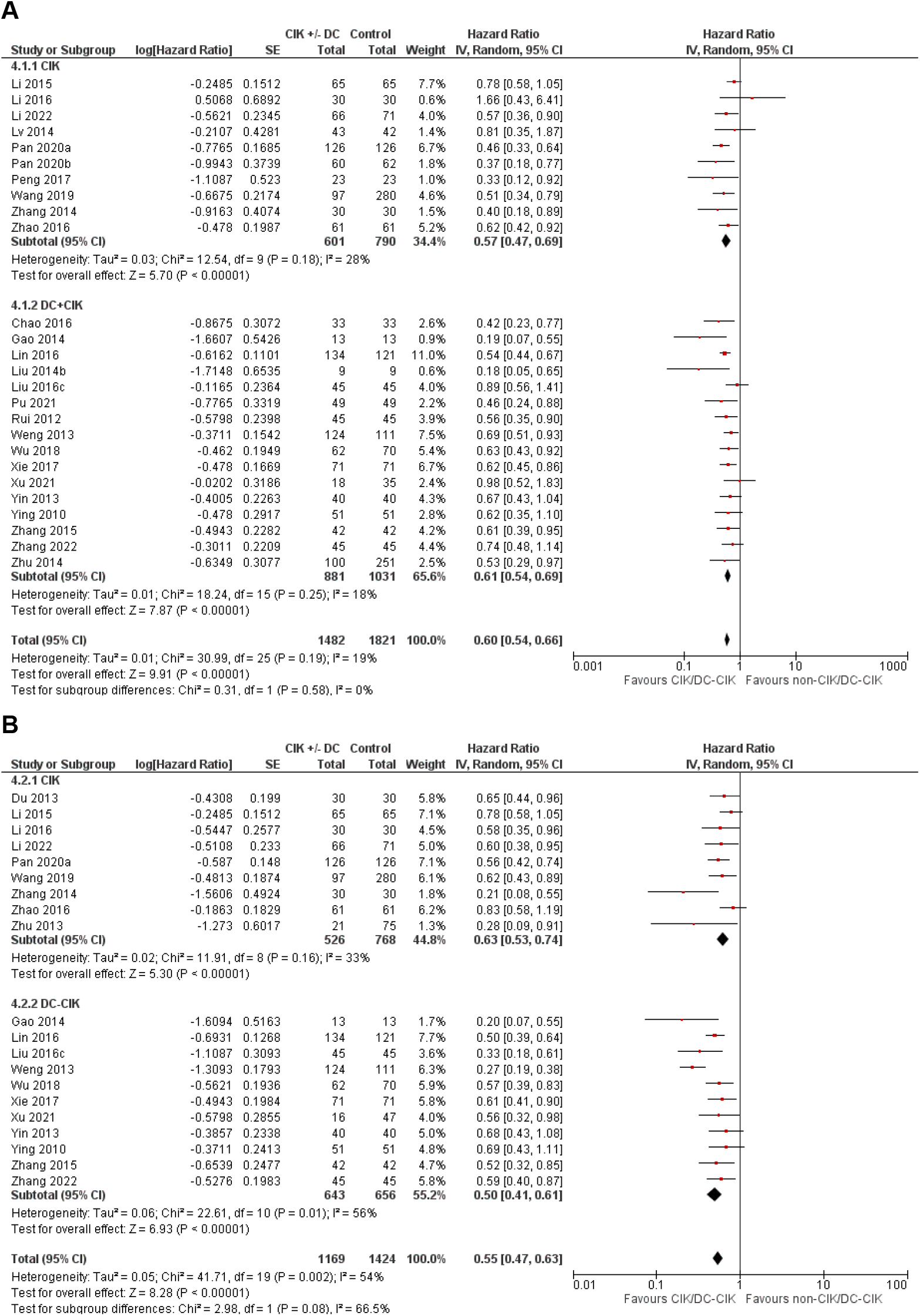
Subgroup analysis by CIK therapy type (with or without DC therapy) for (A) overall survival (OS) and (B) progression-free survival (PFS). Twenty-six studies involving 3,303 patients and twenty studies involving 2,593 patients contributed data to OS and PFS analysis respectively. CIK, cytokine-induced killer cell; DC, dendritic cell.

#### Concurrent CIK/DC-CIK therapy vs sequential CIK/DC-CIK therapy

Subgroup analysis was performed comparing studies where CIK/DC-CIK therapy was administered either concurrently or sequentially with the non-CIK/DC-CIK therapy. For OS analysis, 16 studies^19,27,31,34,36,38,43,58,60,64,65,69,74,76,84,86^ involving 2,000 patients with concurrent administration and 8 studies^17,26,28,29,32,35,68,79^ involving 846 patients with sequential administration were considered (Figure 6A). CIK/DC-CIK therapy administered in either manner improved OS; the HR was 0.63 (95% CI 0.56-071) for concurrent administration and 0.59 (95% CI 0.53-0.65) for sequential administration. A test for subgroup differences reached statistical significance (I^2^=76.3%, p=0.04) with lower HR being observed for sequential administration, although 95% CIs of the 2 subgroups overlapped each other. Subgroup analysis on PFS was similarly in favour of CIK/DC-CIK therapy for both concurrent (HR=0.56, 95% CI 0.46-0.67) and sequential administration (HR=0.54, 95% CI 0.46-0.63) (Figure 6B). Twelve studies^19,25,27,31,34,36,58,64,74,76,84,86^ involving 1460 patients who had concurrent administration and 5 studies^26,28,29,35,79^ involving 580 patients who had sequential administration were evaluated and a test for subgroup differences did not meet statistical significance (I^2^=0%, p=0.43).

**Figure 6.**
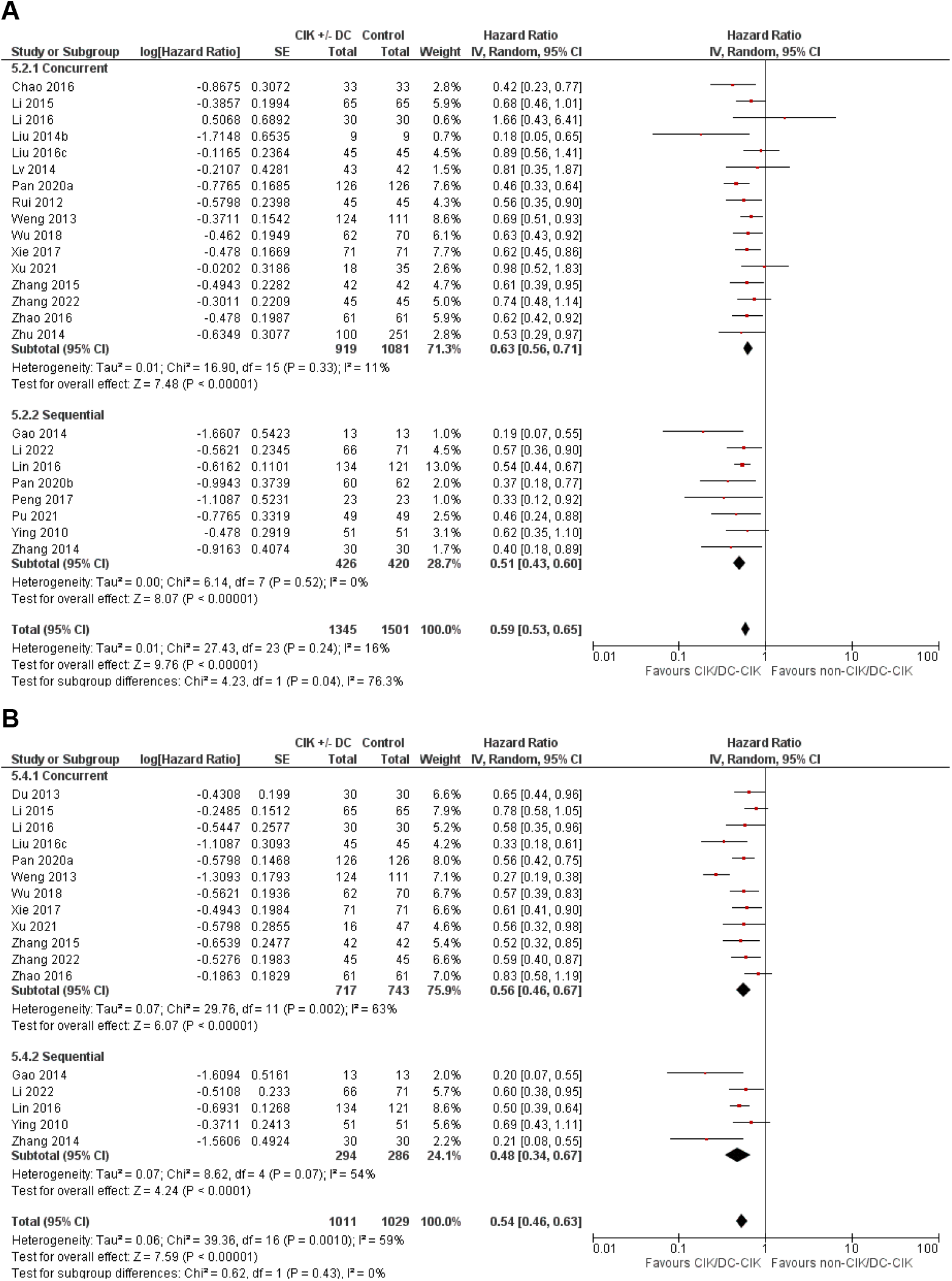
Subgroup analysis by CIK/DC-CIK therapy administration timing for (A) overall survival (OS) and (B) progression-free survival (PFS). Twenty-four studies involving 2,846 patients and seventeen studies involving 2,040 patients contributed data to OS and PFS analysis respectively. CIK, cytokine-induced killer cell; DC, dendritic cell.

## Discussion

Chemotherapy with/without biological therapy remains the standard treatment for CRC patients with high-risk resected disease, and the majority of those with advanced disease. This therapeutic approach is associated with limited survival benefit, unlike immunotherapy, which has demonstrated long-term survival outcome in some solid tumours owing to its mechanism of action^93,94^. New therapeutic approaches which involve modulation of the immune system may provide new treatment options for a broader range of CRC patients, and improve their survival outcome. Autologous adoptive immunotherapy such as CIK therapy represents a highly personalised cancer treatment. While it remains a non-standard treatment option for solid cancers, there are a growing number of clinical trials examining such immunotherapy^95^.

Our study demonstrated that providing CIK or DC-CIK therapy to CRC patients improved OS, PFS and ORR compared standard treatment. The upper 95% CI of pooled HRs for 5-year OS rate and 3- and 5-year PFS rates exceeded 0.85, a commonly applied cut-off to delineate no effect from an important effect, raising the possibility that the observed benefit for these endpoints may not be precise. However, for all the other endpoints, the observed HRs favouring CIK/DC-CIK therapy appeared robust. The OS and PFS benefit of CIK/DC-CIK therapy persisted when prospective randomised studies alone were examined in the subgroup analysis, with no subgroup differences being identified compared to non-randomised studies. While the number of randomised studies assessed was small, HRs and associated 95% CIs reported by each study, especially for the OS endpoint, were all comparable, indicating consistency in the results and so strengthening the overall finding.

Subgroup analysis by CRC disease stage indicated a lack of differences for both OS and PFS. However, the observed 95% CIs associated with the pooled HRs were persistently narrower for Stage IV patients compared to Stage I-III patients, with the upper limits of 95% CIs for Stage I-III patients exceeding 0.85 for both endpoints. Together with the uncertainties around the best way to incorporate CIK/DC-CIK therapy into the established 3-6 months of mono- or doublet-adjuvant chemotherapy, depending on the disease stage and accompanying other prognostic factors, our study highlights that patients with Stage IV disease may be a more suitable target to evaluate CIK/DC-CIK therapy application, at least initially. The immunosuppressive effect of cancer surgery, including T cell and NK cell dysfunction and expansion of myeloid-derived suppressor cells and regulatory T cells in the postoperative period, has been described previously^96^, although how this affects the anti-tumour activity of CIK/DC-CIK therapy is not known.

Subgroup analysis based on combining DC therapy with CIK therapy revealed statistically significant subgroup differences in favour of DC-CIK over CIK therapy for PFS, but not for OS. DCs are major antigen-presenting cells and the essential link between the innate and adaptive immune systems^97^. Co-culturing of CIK cells with DCs results in increased CIK cytolytic function, including cytotoxic activity against a tumour cell line resistant to CIK cells cultured in absence of DCs^14^. This review observed more patients who received DC-CIK therapy than CIK therapy, however, the results suggest that the addition of DC therapy to CIK therapy does not have a strong clinical benefit, as only statistical significance was observed for PFS and not OS. This result points to the need for future clinical trials investigating the benefit of including DC therapy in CIK therapy, and whether other combinations such as immune checkpoint inhibitors or CAR-T incorporation with CIK therapy may be of better value for CRC patients.

Subgroup differences were similarly detected for OS for concurrent versus sequential administration of CIK/DC-CIK. Subgroup analyses for both PFS by CIK therapy type and for OS by CIK therapy administration timing had similar HRs with highly overlapping 95% CIs, making it unclear whether the differences are clinically meaningful. The timing of CIK/DC-CIK delivery for CRC patients may not be critical and could be selected based on logistical issues.

There have been two previous publications that systematically reviewed the literature for CIK/DC-CIK therapy in CRC^16,98^. In 2010, Zhang and Schmidt-Wolf, in cooperation with Stanford University, established the International Registry on CIK Cells (IRCC) to evaluate clinical trials of CIK therapy^98,99^. The registry identifies both prospective and retrospective clinical trials involving CIK therapy for cancer treatment from PubMed, Web of Science Core Collection, WHO International Clinical Trials Registry Platform, ClinicalTrials.gov as well as proceedings of the American Society of Clinical Oncology and European Cancer Conference Annual Scientific Meetings. In addition, the IRCC incorporates clinical trials submitted by individual researchers for inclusion^100^. In 2020 the registry recorded 106 clinical trials, of which only six examined CIK therapy in CRC patients^98^. This contrasts with the twenty-nine trials including 2,610 CRC patients reported in the published systematic review and meta-analysis in 2017 by Zhang *et al*., which purely compared the clinical benefit of CIK therapy plus chemotherapy to CIK therapy in CRC patients with advanced disease^16^. They also utilised two Chinese databases, CNKI and Wanfang Data, in addition to the English databases Cochrane Library, Embase and PubMed. The majority of the studies were published in Chinese similar to our findings.

To date China has taken the lead in research of adoptive immunotherapy including CIK therapy^15,101^. Therefore, the inclusion of articles published in Chinese was necessary to comprehensively review the currently available literature examining clinical efficacy of CIK therapy in CRC. Additionally, the current work included clinical trials which compared CIK therapy to non-CIK treatment not limited to chemotherapy, to increase the number of trials assessed. Consequently, the review considered 70 studies involving 6,743 patients and is the largest systemic review on CIK/DC-CIK therapy in CRC. It meta-analysed OS and PFS, the two most important clinical endpoints in assessing efficacy of any cancer therapy. Endpoints covered by Zhang^16^ were limited to OS and DFS rates as well as ORR. The CRC population covered by this review is also broader having included patients at all stages.

This study has a number of limitations. The heterogeneity observed in the clinical study design requires caution when interpreting results. There are general guidelines for the production of the CIK therapy. The CIK therapy product is generated from PBMCs cultured for 21-28 days in the presence of anti-CD3 stimulation and the cytokines interferon-gamma and interleukin-2. Prior to transfusion, the therapy product is expected to have minimum percentage of NK-like T cells^102^. While having basic production guidelines makes reproducing this therapy achievable, we observed heterogeneity in the culture systems used to generate these cells, including the media, concentration of stimuli and cytokines used, and intervals of cytokine addition in culture. Characterisation of the cell therapy product prior to transfusion to meet the guidelines was normally not provided. Clinical parameters such as anti-cancer treatment history, demographics, and number of treatment cycles were also observed to be heterogenous among the studies analysed. These variables could contribute to the heterogeneity observed in our analysis that was not rectified by our subgroup analyses. As the studies identified were all undertaken in China, clinical trials in non-Chinese ethnicity are needed to confirm its efficacy outside of Chinese patients. Finally, the possibility of publication bias was raised as only a handful of studies reported negative outcomes of CIK/DC-CIK therapy for the efficacy endpoints assessed.

Despite these limitations, our data strongly support that complementing conventional treatment regimens with CIK/DC-CIK therapy in patients with CRC provides clinical benefits. By highlighting the parameters that contribute to the heterogeneity in the study designs, we suggest that standardisation of these will lead to greater adoption of CIK therapy worldwide.

## Conclusion

CIK therapy in combination with standard treatments, in particular chemotherapy, provides clinical benefit for CRC patients. The benefit existed whether the included studies were prospective and randomised or not, strengthening the finding. CIK therapy was well tolerated, with fever being the most common adverse event. While DC therapy is commonly combined with CIK therapy for CRC patients, our study suggests that this may not provide extra benefit. The findings support further evaluation of the clinical utility of CIK therapy in CRC.

## Data Availability

All data produced in the present work are contained in the manuscript

## Declarations

### Contributors

Conception Design: CMYL, YT and KF. Collection and/or assembly of data: CMYL, YT, RL, and JL. Data analysis and interpretation: CMYL, YT, BD, ES and KF. Manuscript Writing: CMYL, YT, ES, PD, and KF. Final Approval of Manuscript: CMYL, YT, BD, RL, JL, PD, ES, TP, GM, and KF.

### Funding

This work was supported by The Hospital Research Foundation (THRF)/Cancer Council SA Beat Cancer Hospital Research Package (GM) and a Tour de Cure Early Career Grant (KF). BD was supported by a Schlumberger Foundation Faculty For Future Fellowship and KF was supported by a THRF Early Career Fellowship.

### Competing Interests

The authors have no relevant financial or non-financial interests to disclose.

### Data Availability

All data generated or analysed during this study are included in this published article and its supplementary material. The datasets used and/or analysed during the current study are available from the corresponding author on reasonable request.

## Notes

### Competing Interest Statement

The authors have declared no competing interest.

### Funding Statement

This work was supported by a The Hospital Research Foundation (THRF)/Cancer Council SA Beat Cancer Hospital Research Package (GM) and a Tour de Cure Early Career Grant (KF). BD was supported by a Schlumberger Foundation Faculty For Future Fellowship and and KF was supported by a THRF Early Career Fellowship.

### Author Declarations

This is a systematic review and meta-analysis. The data are derived from publicly available journal articles.

