## Supplementary File for "Use of cytokine-induced killer cell therapy in colorectal cancer patients: a systematic review and meta-analysis"

*Supplementary Data*

### Supplementary Tables

**Supplementary Table 1.** Search strategy for Embase

#1 Cytokine-Induced Killer Cells/
#2 cytokine induced killer cell*.mp. [mp=title, abstract, original title, name of substance word, subject heading word, floating sub-heading word, keyword heading word, organism supplementary concept word, protocol supplementary concept word, rare disease supplementary concept word, unique identifier, synonyms]
#3 (cytokine induced adj6 killer cell*).mp.  [mp=title, abstract, heading word, drug trade name, original title, device manufacturer, drug manufacturer, device trade name, keyword heading word, floating subheading word, candidate term word]
#4 2 or 3
#5 1 or 4
#6 *colorectal cancer*/ or colorectal neoplasm*.mp. or colon cancer*.mp.  [mp=title, abstract, heading word, drug trade name, original title, device manufacturer, drug manufacturer, device trade name, keyword heading word, floating subheading word, candidate term word]
#7 exp colon tumor/
#8 exp rectum tumor/
#9 6 or 7 or 8
#10 5 or 9

**Supplementary Table 2.** Search strategy for MEDLINE

#1 Cytokine-Induced Killer Cells/
#2 cytokine induced killer cell*.mp. [mp=title, abstract, original title, name of substance word, subject heading word, floating sub-heading word, keyword heading word, organism supplementary concept word, protocol
#3 (cytokine induced adj6 killer cell*).mp. [mp=title, abstract, original title, name of substance word, subject heading word, floating sub-heading word, keyword heading word, organism supplementary concept word, protocol supplementary concept word, rare disease supplementary concept word, unique identifier, synonyms]
#4 2 or 3
#5 1 or 4
#6 exp Colorectal Neoplasms/
#7 *colorectal cancer*/ or colorectal neoplasm*.mp. or colon cancer*.mp. [mp=title, abstract, original title, name of substance word, subject heading word, floating sub-heading word, keyword heading word, organism supplementary concept word, protocol supplementary concept word, rare disease supplementary concept word, unique identifier, synonyms]
#8 6 or 7
#9 5 and 8

### Supplementary Figures

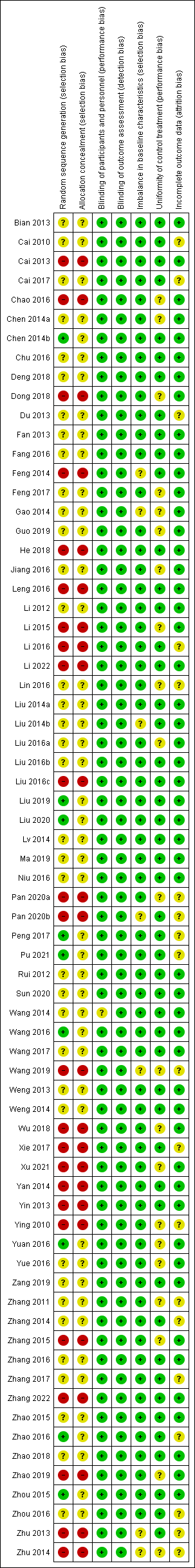

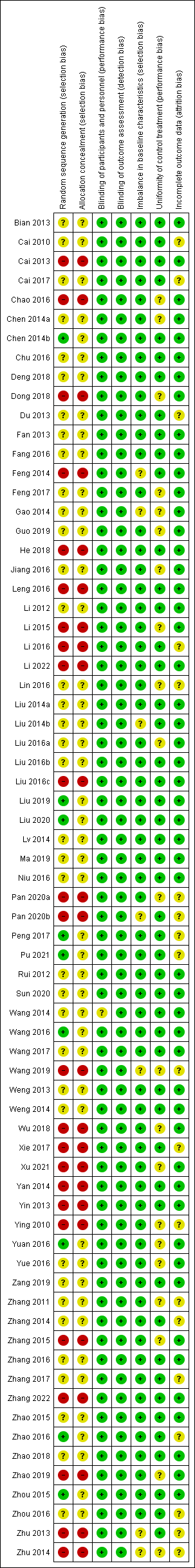

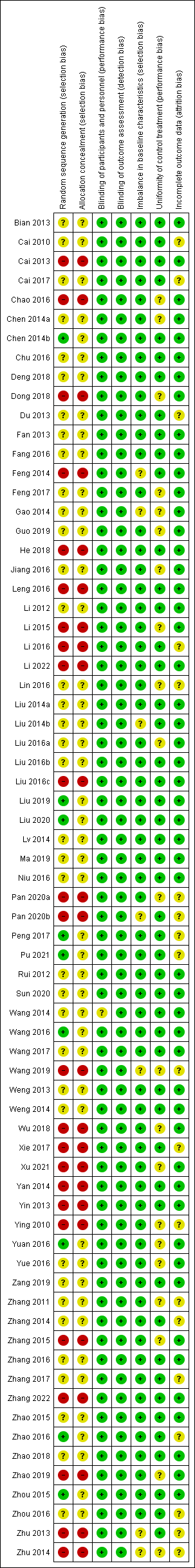

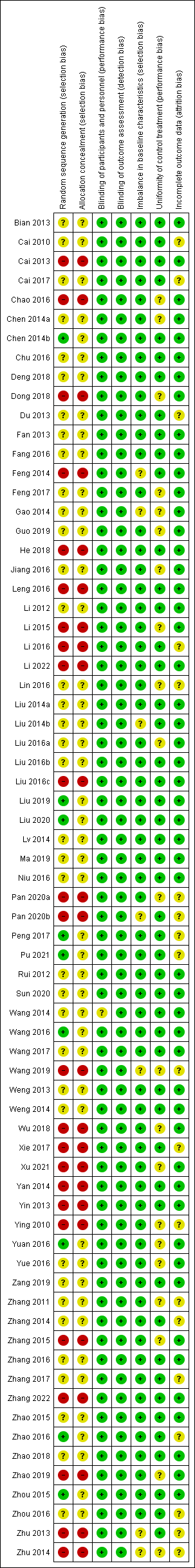

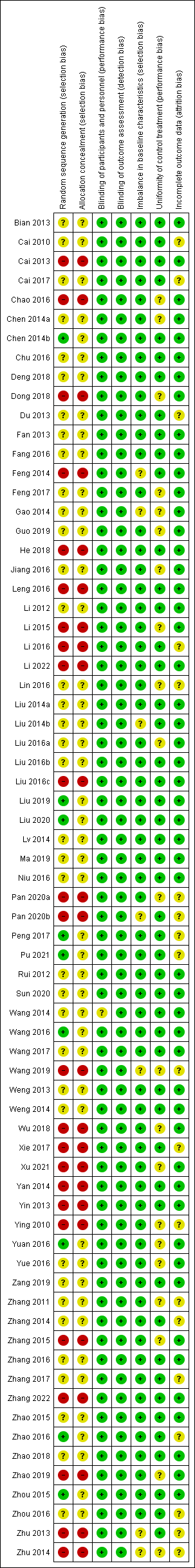

**Supplementary Figure 1.** Risk of bias assessment summary

**A**

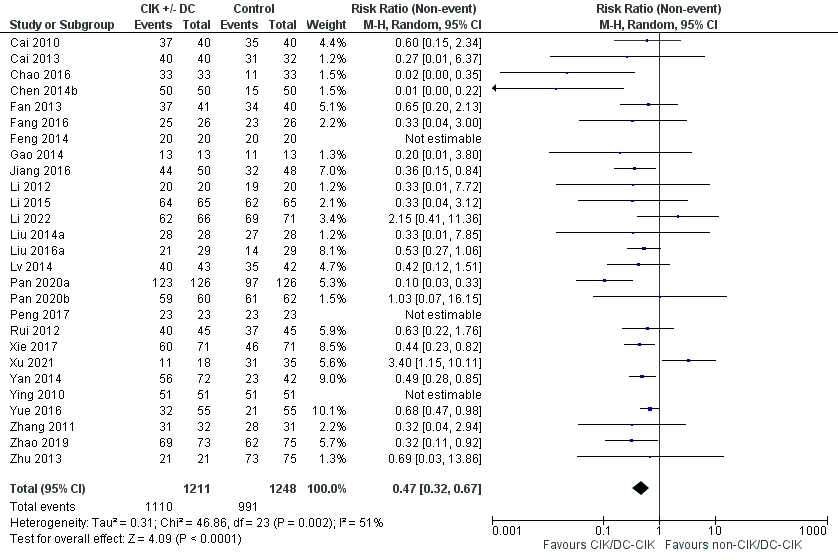

**B**

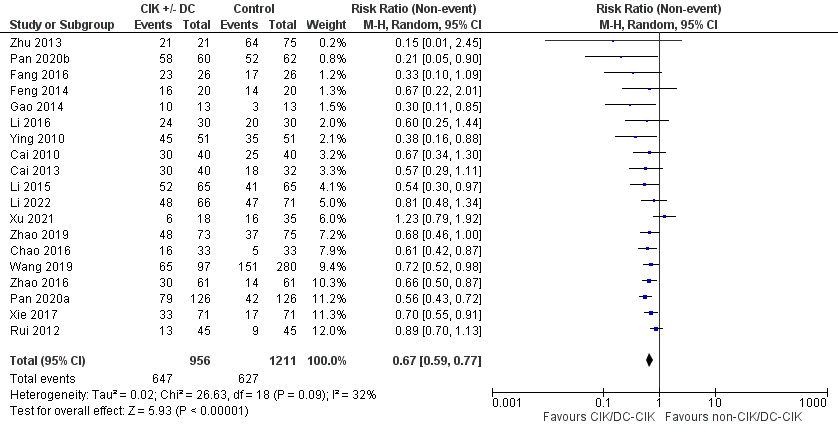

**C**

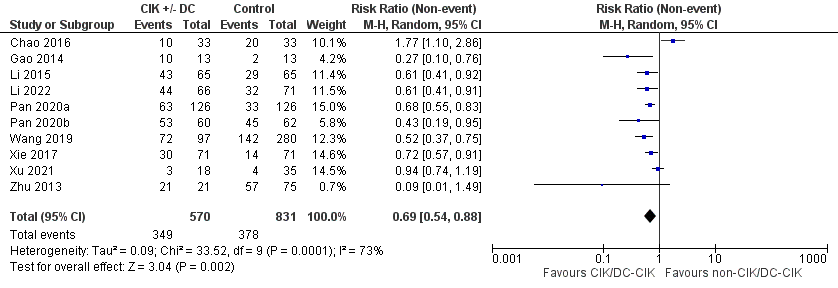

**Supplementary Figure 2.** CIK/DC-CIK therapy versus non-CIK/DC-CIK therapy for (A) 1-year, (B) 3-year and (C) 5-year overall survival (OS) rates. Twenty-seven studies involving 2,459 patients, nineteen studies involving 2,167 patients and ten studies involving 1,401 patients contributed data to 1-, 3- and 5-year OS rate analysis respectively. CIK, cytokine-induced killer cell; DC, dendritic cell.

**A**

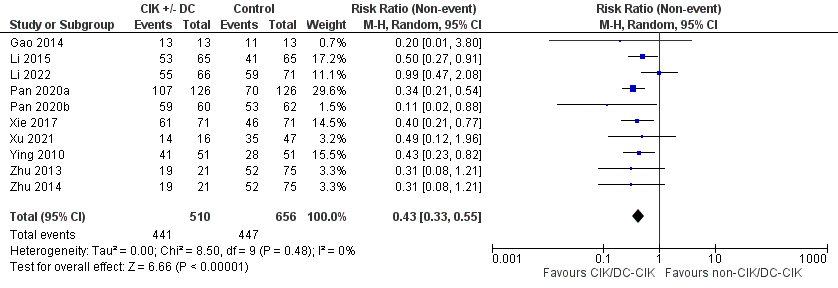

**B**

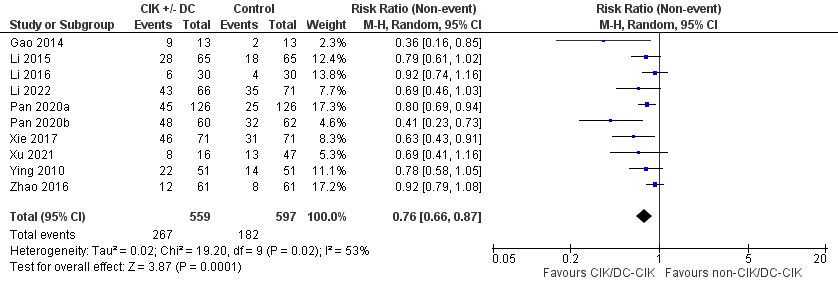

**C**

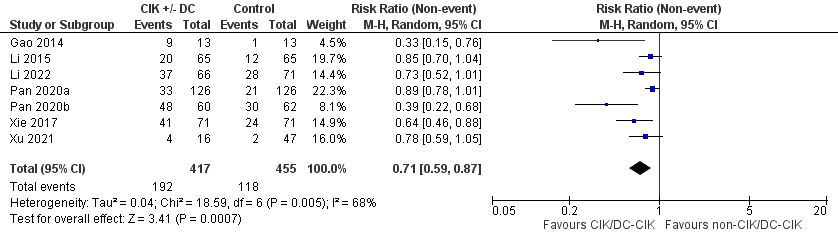

**Supplementary Figure 3.** CIK/DC-CIK therapy versus non-CIK/DC-CIK therapy for (A) 1-year, (B) 3-year and (C) 5-year progression-free survival (PFS) rates. Ten studies involving 1,166 patients, ten studies involving 1,156 patients and seven studies involving 872 patients contributed data to 1-, 3- and 5-year PFS rate analysis respectively. CIK, cytokine-induced killer cell; DC, dendritic cell.

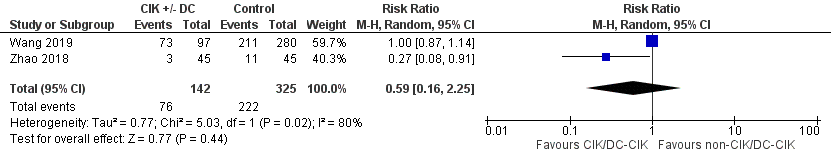

**Supplementary Figure 4.** CIK/DC-CIK therapy versus non-CIK/DC-CIK therapy on any adverse events. Two studies involving 467 patients contributed data to any adverse events analysis. CIK, cytokine-induced killer cell; DC, dendritic cell.

**A**

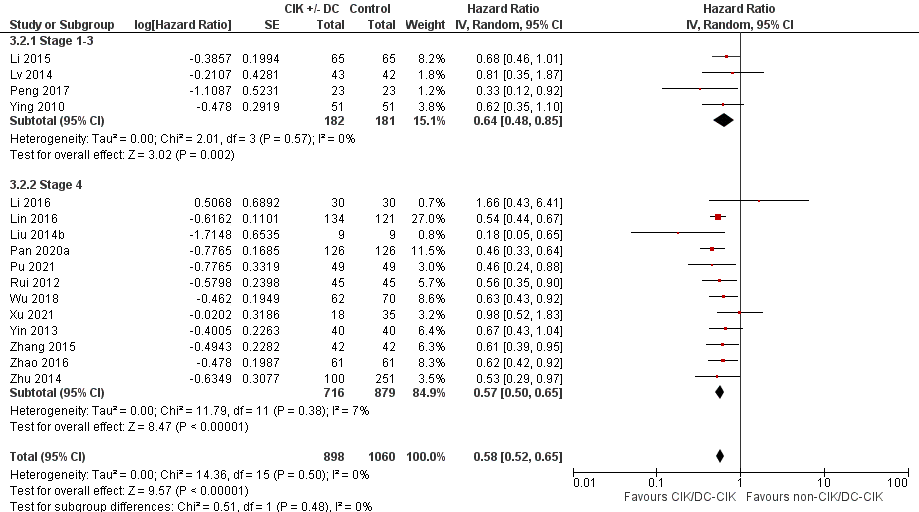

**B**

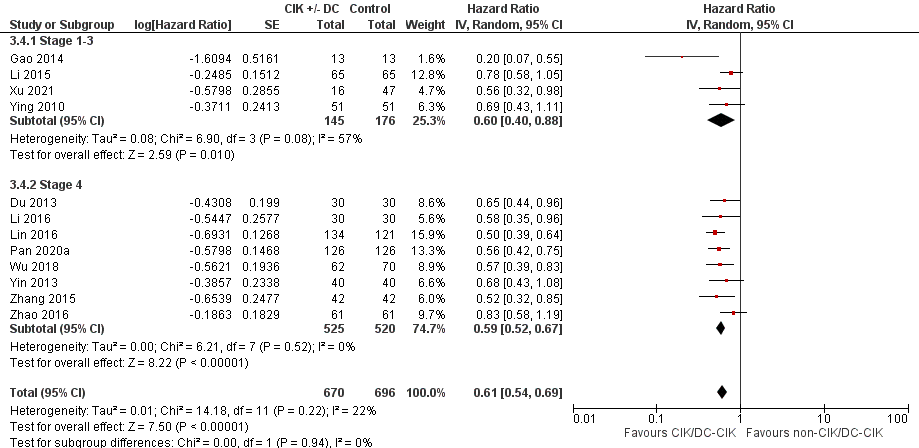

**Supplementary Figure 5.** Subgroup analysis by disease stage for (A) overall survival (OS) and (B) progression-free survival (PFS) between CIK/DC-CIK therapy versus non-CIK/DC-CIK therapy. Sixteen studies involving 1,958 patients and twelve studies involving 1,366 patients contributed data to OS and PFS analysis respectively. CIK, cytokine-induced killer cell; DC, dendritic cell.
